# BACTERIAL INFECTION AND ANTIBIOTIC RESISTANCE IN PATIENTS WITH HUMAN IMMUNODEFICIENCY VIRUS PRESENTING LOWER RESPIRATORY TRACT INFECTION IN NIGERIA

**DOI:** 10.1101/2021.09.05.21263138

**Authors:** Joel Iko-ojo Oguche, Rebecca Olajumoke Bolaji, Josiah Ademola Onaolapo, Samuel Eneojo Abah, Vivian Gga Kwaghe, Shedrack Egbunu Akor, Sarah Shaibu, Tijani Salami, Gloria Eleojo Eneojo-Abah

## Abstract

Lower Respiratory Tract Infection (LRTI) is associated with mortality and prolonged antibiotics use among HIV/AIDS patients. Sputum samples were collected from 134 HIV/AIDS patients with LRTI and productive cough, visiting University of Abuja Teaching Hospital, Nigeria, to determine the bacterial aetiologies and antibiotic resistance profile. Adequate for culture sputum samples were observed in only 119 subjects (75 females and 44 males) and cultured using standard procedure. Isolates were identified by biochemical method and 16SrRNA gene amplification, purification, sequencing and database nucleotide blast. Antibiotic susceptibility tests were performed and interpreted according to Clinical and Laboratory Standard Institute (CLSI) procedures. Bacteria associated LRTI was found in 20.2% of the patients and Klebsiella *pneumonia* (29.7%); *Enterobacter cloacae* (16.7%); *Enterobacter hormaechei* subsp. *xiangfangensis* (12.5%); *Pseudomonas parafulva*; *Pseudomonas aeruginosa*; *Pseudomonas alcaliphila; Klebsiella aerogenes* (8.3%); *Comamonas testosteroni*; *Escherichia coli*; *Acinetobacter junii*; *Acinetobacter soli* and *Acinetobacter baumannii* were implicated. Isolates show high resistance to amoxicillin-clavulanic acid at 94.1%, trimethoprim-sulfamethoxazole at 75.0% and cefotaxime at 70.0%. The Multiple Antibiotic Resistance (MAR) index of most isolates (62.5%) is within 0.3-0.8 in a range of 0.0 to 0.8. Isolates of the same species were found to have different MAR index in different patients. However, *E. cloacae, E. hormaechei* subsp. *xiangfangensis, A. baumanni* and 71.4% of *K. pneumonia* were Multi-drug Resistant (MDR). Interestingly, Gentamycin, Ciprofloxacin and Imipenem were effective against MDR isolates and showed significant negative correlation with MAR index. We propose that antibiotics with efficacy against MDR isolates could be used to down regulate the selective pressure of other antibiotics, and these MDR pathogens, including those rarely associated with human infection poses potential threat, similar to Methicillin Resistant *Staphylococcus aureus* (MRSA). Particularly, among the immunocompromised. Furthermore, HIV/AIDS patients present good metrics for profiling the burden and spread of antibiotic resistant bacteria in poor countries.

## 1.0 INTRODUCTION

HIV/AIDS patients by far are the most at risk and the most affected by lower respiratory tract infections (LRTI) in the human population. LRTI is 25-fold more common in patients with HIV than in the general community, occurring in up to 90 cases per 1,000 persons yearly (1). Pulmonary infections are a leading cause of morbidity, hospitalisation and mortality in HIV-infected individuals (2). Despite the use of highly active antiretroviral therapy (HAART) which has led to a remarkable improvement in the immune status of HIV/AIDS patients, pulmonary infections continue to be a devastating disease in HIV/AIDS patients than in HIV-negative individuals (3).

Since the description of HIV/AIDS in 1981 (4), the lungs has been the site most often affected by the HIV disease (5). This is partly due to impaired immune function as a result of depletion of CD4^+^ cells and being externally exposed to aerosolized antigens and microorganisms (6).

In developing countries like Nigeria, poor sanitation, poor infection management, economic constraints, limited understanding of HIV transmission, high-risk sexual behaviour (sometimes in conjunction with intravenous drug use), and inconsistent access to highly active antiretroviral therapy (HAART) once seropositive, currently place developing nations in danger of increasing HIV/AIDS prevalence and continuous burden on antibiotics (7).

HIV linked antibiotic therapy is a factor for increased antibiotic pressure. The practice of frequent and prolonged use of antibiotics for treatment and or prophylaxis due to increased risk to opportunistic infections because of AIDS-immunodeficiency is a potential for the development of multidrug-resistant bacteria. Even when appropriately used, it has been observed that constant use of antibiotics advances the emergence and maintenance of drug-resistant microbes because it exerts a strong selective pressure upon drug-sensitive pathogens (8,9). Antibiotic abuse may be on-going too within the HIV/AIDS population as their fragile health condition can influence indiscriminate prescription and self-medication. HIV/AIDS is therefore a risk factor for bacterial infection (3). The number of infections caused by multidrug-resistant bacteria is increasing and poses a challenge among healthcare workers on the continued ineffectiveness of antibiotics in the management of HIV/AIDS patients (10,11).

The use of antibiotic prophylaxis is known to create an evolutionary trade-off, wherein despite the potential improvement in host health, a fitness benefit is conferred to resistant bacterial strains. However, within the context of an HIV/AIDS-prevalent host pool, the elevation of resistant strain fitness that arises as an inevitable by-product of prophylaxis use may represent a greater health risk—especially given the speed with which resistant bacterial strains become dominant (12). Studies have shown association between long term use of trimethoprim-sulfamethoxazole with increase in development of resistance to trimethoprim-sulfamethoxazole among bacteria in HIV/AIDS patients (12,13).

It is important therefore to continuously mount antimicrobial surveillance study on HIV/AIDS patients across different setting in order to monitor trends in bacterial antibiotic resistance and to also identify emerging bacterial aetiologies of LRTI, particularly in developing countries where the burden of respiratory tract infection is highest (14); only a small proportion of poor quality antimicrobials are available, which may be sold over the counter without proper diagnostic guidance (15); and inadequate hygiene and infection control in hospitals may increase the spread of multidrug resistant pathogens (16). The information will enable effective public health intervention and guide physicians on the choice of antibiotics in the empirical management of LRTI in HIV/AIDS patients.

The vulnerability of the immunocompromised, like the HIV/AIDS to many opportunistic pathogens makes them a good metrics for monitoring the spread of antibiotics resistance and the need to continuously mount surveillance on the aetiologies of bacterial infection and resistance to antibiotic from a One health approach (17,18). Hence, in this study, HIV/AIDS patients visiting out-patient clinics and presenting with LRTI at the University of Abuja teaching Hospital were recruited to determine the antibiotic resistance pattern of bacterial isolates associated with LRTI.

## 2.0 METHOD

### Ethic Approval/Informed Consent

The ethic approval for this study was obtained from the Health Research Ethics Committee (HREC) of the University of Abuja Teaching Hospital. Informed consent was also obtained from patients before being enrolled for this study in accordance with the World Medical Association ethical principle for research involving human.

### Study site

This study was performed in the University of Abuja Teaching Hospital (UATH) Gwagwalada, Federal Capital Territory a 350-bed hospital. Abuja is situated in the centre of Nigeria and it’s the capital of Nigeria. The FCT has a very diverse population in terms of socioeconomic status. Those with a high standard of living are majorly concentrated in Abuja (the central area) of FCT and those with low socioeconomic status are concentrated in a densely populated suburbs around the city. Being the seat of power, prostitution could be very rampant with HIV prevalence rate of about 1.5% that is presently above the national average and the highest in the Middle Belt Region of Nigeria (MBRN).

### Sputum collection and bacterial isolation

A total of 134 patients with lower respiratory tract infection were recruited between May and September 2017, out of which 119 patients were able to produce sputum that met standard criteria for sputum culture. All sputum samples were collected aseptically in a sterile screw cap container and transported to the laboratory for culturing within 1 hour after samples collection and were processed in accordance to the standard protocol (19). Sputum samples were cultured using Blood, Chocolate, and MacConkey agars (Sigma-Aldrich) and the manufacturers’ instructions. Briefly, a sterile calibrated wire loop was used to streak an aliquot of the sputum on the agar plate and were grown under different conditions, at 37°C for 24 hours on Chocolate agar plates in an anaerobic condition; aerobically on MacConkey agar and blood agar plates at 37°C for 24 hours (10,20). Discrete colony growing along the streak pattern were sub-cultured on nutrient agar slant and stored at 4°C.

### Antibiotic Susceptibility Test

The antibiotic susceptibility testing of isolates were determined as previously described (21). Briefly, a discrete colony suspension equivalent to 0.5 McFarland standard and containing approximately 1.5 × 10^8^ cfu/ml) were inoculated on Mueller Hinton agar (Oxoid) using a sterile disposable spreader. Antibiotics of varying concentration disc were placed on the agar surface on which the bacteria were inoculated using the spread plate technique. The plates were incubated at 37°C for 18h. After incubation, the plates were examined and the zones of inhibitions were measured with the aid of a venier caliper (0.01mm sensitivity). The antibiotic discs that was used contain 10µg of Gentamycin; 30µg Cefoxitin; 5µg Ciprofloxacin; 1.25µg+23.75µg Trimethoprim-sulfamethoxazole; 30µg Tetracycline; 10µg Streptomycin; 30µg Ceftazidime; 30µg Cefotaxime; 20µg+10µg Amoxicillin-Clavulanic Acid; 10 µg Imipenem (Oxiod) and were selected according to range of locally available antimicrobials, local prescribing policies and Clinical and Laboratory Standard Institute (CLSI) recommendation (22). Results were interpreted according to CLSI (2017) and the European Committee of Antimicrobial Susceptibility Testing (EUCAST), 2019 (22,23).

### Gram stain and Biochemical analysis

Gram staining was carried out to determine the morphology of the organisms according to standard protocol (19). Oxidase, Citrate, Methyl red and Voges-Proskauer test (MR-VP), Triple Sugar Iron (TSI) test and Cetrimide utilization test were used for preliminary identification of Gram negative bacteria (19,24), data not shown.

### Genomic DNA isolation

Genomic DNA was isolated using GeneJET DNA purification Kit (Thermoscientific) and the manufacturer’s instructions. The quality and amount of genomic DNA was accessed using Nanodrop (Thermoscientific) with 1µl of the DNA sample. The concentration of DNA was measured in µg/mL, data not shown.

### Amplification of 16SrRNA genes

Genomic DNA and specific primers covering both variable and constant regions of 16SrRNA gene, 8F (5’-AGA GTT TGA TCC TGG CTC AG-3’) and U1492R (5’-GGT TAC CTT GTT ACG ACT T-3’) producing a product length of about 1484bp, approximately 1.5kb (Figure 1) were used for this study to amplify the 16SrRNA genes. The PCR was performed with a GoTaq Green Master Mix (Promega) and the manufacturer’s instruction. The PCR reaction include an initial denaturation at 95°C for 3 minutes and 35cycles of denaturation steps at 98°C for 30 seconds; 60.8°C, 30 second annealing and 72°C for 1 minute extension. An additional final extension was performed at 72°C for 1 minute as the final step.

**Figure 1:**
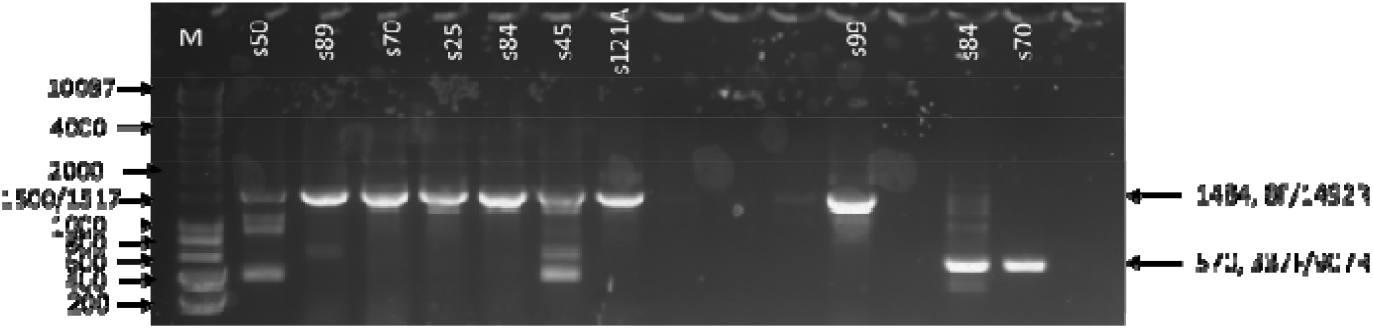
Agarose Gel Photo showing PCR product of the right size. M = Molecular ladder (1kb hyper ladder; Bioline); s50, s70 and similar designations are isolates IDs: numbers represent band sizes of PCR products. 33F/907R are other 16SrRNA primers not used in this study. The right product size in the figure is 1484bp, produced by 8F/1492F primer pair.

**Figure 2:**
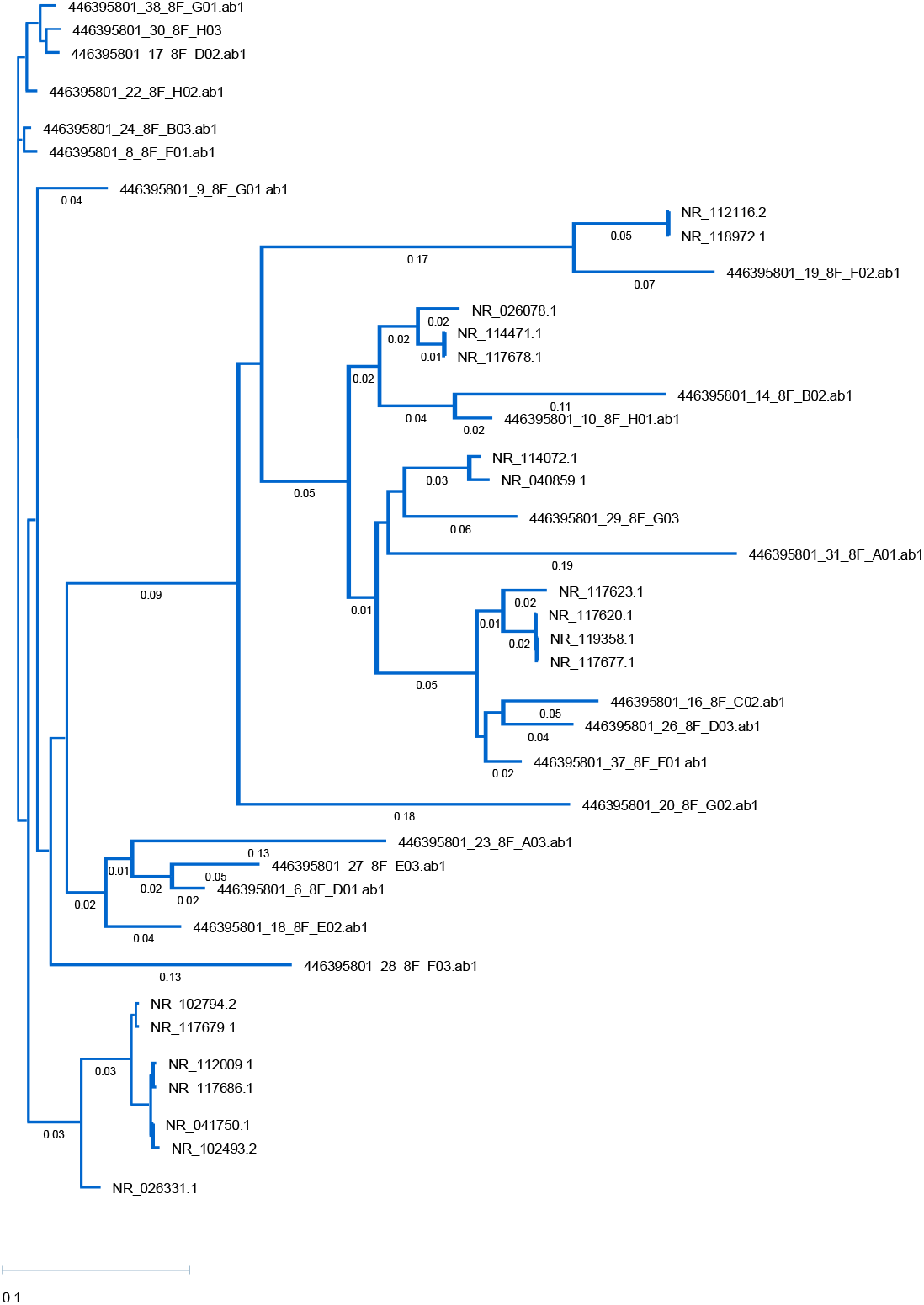
Phylogenetic tree based on sequenced 16SrRNA gene. The sequence was blasted to identify the organisms. The divergence of the organism against a given sequence is indicated in the tree. The tree shows how the sequenced 16SrRNA is very similar (based on low divergence bootstrap) to the bacteria identified based on Max. Score and E values. This indicated that the 16SrRNA gene sequence is specific to identifying our bacterial isolates, with only 0.01-0.05 divergence on the average. NR_118568.1 = *Enterobacter cloacae* strain ATCC 13047; NR_26208.1= *Enterobacter hormaechei* subsp. *xiangfangensis* 10-17; NR_041750.1= *Klebsiella pneumoniae* subsp. ozaenae strain ATCC 11296; NR_117686.1= *Klebsiella peumoniae* strain DSM 30104; NR_102493.2= *Klebsiella aerogenes* strain KCTC 2190; NR_117623.1= *Acinetobacter junii* strain ATCC 17908; NR_044454.1= *Acinetobacter soli* strain B1; NR_117677.1= *Acinetobacter baumanni* strain DSM 30007; NR_040859.1= *Pseudomonas parafulvai* strain DSM17004; NR_029161.2= *Comamonas testosterone* strain KS0043; NR_114042.1= *Escherichia coli* strain NRBC 102203; AP_019665.1= *Klebsiella pneumoniae* strain TA6363; NR_117678.1= *Pseudomonas aeruginosa* DSM50071; NR_114072= *Pseudomonas alcaliphila* strain NRBC 102411; 446395801_18_8F_E02.abi and similar designations are sequence ID of isolates.

### Verification of PCR products

The PCR products were checked by agarose gel electrophoresis. A 0.7-1% molecular grade (DNase/RNase free) agarose (Fisher Bioreagents) and TAE buffer was used. Briefly, 0.7g of agarose was measured on a weighing balance (OHAUS) after being zeroed to remove the tare weight and was dissolved in 100ml, 1x TAE buffer made from 10x TAE stock (48.4g Trise base; 11.4mL glacial acetic acid; 17.4M and 3.7g EDTA). The agarose solution was melted in microwave and 1 ul of Sybr^®^safe DNA gel stain was added prior to casting the gel. The same 1x TAE buffer (1 litre) with an aliquot of Sybr^®^safe DNA gel stain was used as the running buffer. A 5µl of DNA ladder (1kb hyperladder; Bioline) and 10µl of PCR product were loaded into each well. The electrophoresis was performed at 100V, 400mA for 50-60 minutes. Gel was visualised for the right product size (band) and documented using a gel documentation apparatus.

### Gel extraction of PCR product and purification

Bands from agarose gel were excised under mild UV light into a 1.5ml Eppendorf tube in a very quick procedure. A QIAquick PCR purification kit (28506; Qiagen) and the manufacturer’s procedure were used to purify the DNA. The QIAquick kit contains a silica membrane assembly for binding of DNA in high-salt buffer and elution with low-salt buffer or water. This purification procedure removes primers, nucleotides, enzymes, mineral oil, salts, agarose, Sybr®safe, and other impurities from DNA samples. Up to 25µl DNA were eluted with the QIAquick purification kit.

### Gene Sequencing

A 10ng/µl of purified PCR product and 5µl of 3.2pmol/µl primers were sent to Source Bioscience for sequencing. Sequence data were received in ‘.seq’ (a text file in fasta format) and ‘.abi’ format. The DNA sequence and the electropherogram were used for further downstream analysis.

### Bacterial identification based on the 16SrRNA gene sequencing

16SrRNA gene sequence, SnapGene^R^ and the NCBI database were used to identify the bacteria. Briefly, the ‘.abi’ sequence data was opened with SnapGene^R^ for visualisation. For the identification of the pathogen, sequences were copied from SnapGene and blasted in the NCBI nucleotide blast (https://blast.ncbi.nlm.nih.gov/Blast.cgi), specifying 16SrRNA sequences for bacteria and archaea databases and optimising the programme for highly similar sequences mega blast. Parameters like the Max. score; Total score; Query cover; E. value; % identity and Accession number were all used to identify the organism.

### Statistical Analysis

We used a MegAlign pro (DNASTAR) to create a divergence tree between the 16SrRNA gene of sequenced isolates and related 16SrRNA genes in the database. The tree shows bootstrap value based on divergence (small divergence, 0.02 bootstrap value means high similarity, 99.98%) between the 16SRNA gene sequence of isolates and other 16SrRNA gene sequence in the NCBI database. A high similarity values based on E-value, Max score, total score, Query cover and % identity and as represented by very low divergence bootstrap value on tree is used to identify the isolates.

MAR index was computed as the ratio of the number of antibiotic resistance of isolates to the total number of antibiotics to which the isolates were exposed. A Pearson correlation and regression analysis (PRISM Software) was used to analyse the resistance and susceptibility pattern of isolates to each of the antibiotics with the MAR index of each isolate (Table 4). The percentage resistance of a given group of isolates to antibiotics were computed (Table 5). The percentage of MDR isolates were computed from the number of isolates groups that showed resistance to more than two antibiotic categories (Table 6).

## 3.0 RESULT

### Clinical demographics

The patient group are within the age of 25-60 and are male and female with and without employment (Table 1). The HIV infection duration ranges from 1-13 years. All patients are on antiretroviral therapy and only 3 of the patients were on the 2nd regimen of antiretroviral therapy (Table 1). Out of 119 patients from whom adequate for culture sputum samples were collected, only 24 (20.2%) had definitive diagnosis for Bacterial Lower Respiratory Tract Infection, BLRTI (Table 2), which are reported in this study. All patients were given cotrimoxazole (Trimethoprim-sulfamethoxazole) prophylaxis (Table 1). The prevalence of bacterial infection in male and female were 27.3% and 16.0% respectively. The prevalence of bacterial infection differs with age group and are 24.5%, 16.9% and 20.0% in 40-59, 20-39 and in >60 years respectively, and with occupational status and are 22.5% and 20.0% in employed and unemployed participants respectively. A prevalence pattern is also observed with patients’ CD4 cell count. Patients with none (CD4 counts of ≥500 cell/mm^3^); mild (CD4 count range from 351-499 cell/mm^3^); advanced (CD4 count range from 201-350 cell/mm^3^) and severe (CD4 counts ≤ 200 cell/mm^3^) immunodeficiencies have infection prevalence of 9.8%, 33.3%, 25.0%, and 18.2% respectively (**Table 1 and 7**). Both the age and the duration of the infection does not correlate with the CD4 counts of the patients as at the time of sputum sample collection. The Age of the study population correlates with duration of the infection and resistance to CTX antibiotics (Tables 1 and 4)

### Rarely pathogenic bacteria are associated with Lower Respiratory Tract Infection in HIV/AIDS patients

Bacteria not reported to have been previously associated with Lower Respiratory Tract Infection (LRTI) and rarely pathogenic bacteria, were identified (Table 2). For example, *Acinobacter soli; Enterobacter hormaechei* subsp *xianfangensis*; *Acinobacter Junii*; *Pseudomonas parafulva, Comamonas testosteroni* and *Psudomonas alcaliphila* (Table 2). These bacteria are associated with plants, environment, nosocomial infection and some are normal flora. Other common LRTI bacteria identified are *Enterobacter cloacae*; *Acinobacter baumannii*; *Klebsiella pneumoniae; Klebsiella aerogenes* and *Pseudomonas aeruginosa* (Table 2).

### Antibiotics susceptibility Test

Antibiotic susceptibility test showed that the percentage resistance varies from 14.3% to 100%. Most resistance were to Amoxicillin-clavulanic acid (AMC); Trimethoprim-sulfamethoxazole (SXT) and Cefotaxime (CTX). *Enterobacter* spp. and *K. pneumoniae* have the highest resistance. All *Enterobacter* spp. were resistant to AMC, SXT, CTX and FOX (Table 5). All *K. pneumoniae* were resistant to AMC and SXT (Table 5). Antibiotics susceptibility tests were designated as resistant, intermediate resistant, susceptible and non-determinate (S102, *Comamonas testosteroni*) based on standard interpretation (Table 3).

**Table 3:** Antibiotics resistance map and MAR Index of isolates associated with BLRTI in HIV/AIDS

| ISOLATE IDs | BACTERIA | CN | SXT | TE | FOX | AMC | CIP | STR | CTX | CAZ | IPM | MAR Index |
| --- | --- | --- | --- | --- | --- | --- | --- | --- | --- | --- | --- | --- |
| S25 | *Klebsiella pneumoniae* strain DSM 30104 | R | R | S | S | R | I | S | S | S | S | 0.3 |
| S46 | *Enterobacter cloacae* strain ATCC 13047 | R | R | R | R | R | I | R | R | R | S | 0.8 |
| S49 | *Enterobacter hormaechei* subsp. *xianfangensis* 10-17 | R | R | S | R | R | I | R | R | R | I | 0.7 |
| S50 | *Enterobacter hormaechei* subsp. *xianfangensis* 10-17 | R | R | R | R | R | I | R | R | R | S | 0.8 |
| S54 | *Enterobacter cloacae* strain ATCC 13047 | R | R | S | R | R | S | R | R | R | S | 0.7 |
| S55 | *Enterobacter cloacae* strain ATCC 13047 | R | R | S | R | R | I | R | R | R | S | 0.7 |
| S59 | *Klebsiella pneumoniae* subsp. *ozaenae* strain ATCC 11296 | S | R | R | R | R | I | R | R | I | S | 0.6 |
| S67 | *Klebsiella pneumoniae* strain DSM 30104 | S | R | R | S | R | R | S | R | I | S | 0.5 |
| S70 | *Klebsiella aerogenes* strain KCTC 2190 | S | S | S | S | S | S | S | I | S | S | 0 |
| S82 | *Klebsiella pneumoniae* strain DSM 30104 | S | R | R | S | R | S | I | S | S | S | 0.3 |
| S83 | *Klebsiella pneumoniae* strain DSM 30104 | S | R | S | S | R | S | S | I | S | S | 0.2 |
| S84 | *Klebsiella aerogenes* strain KCTC 2190 | S | S | S | S | R | S | S | S | S | S | 0.1 |
| S86 | *Acinetobacter junii* strain ATCC 17908 | S | I | S | nd | nd | S | nd | R | S | S | 0.1 |
| S87 | *Acinetobacter baumannii* strain DSM 30007 | S | R | I | nd | nd | R | nd | R | S | S | 0.4 |
| S89 | *Enterobacter hormaechei* subsp. *xianfangensis* 10-17 | S | R | R | S | R | S | R | R | S | S | 0.5 |
| S90m | *Acinetobacter soli* strain B1 | S | S | S | nd | nd | R | nd | I | S | S | 0.14 |
| S99 | *Pseudomonas parafulvai* strain DSM 17004 | S | nd | nd | nd | nd | R | nd | nd | nd | S | 0.3 |
| S102 | *Comamonas testosteroni* | nd | nd | nd | nd | nd | nd | nd | nd | nd | nd | - |
| S104 | *Klebsiella pneumoniae* strain DSM 30104 | S | R | R | S | R | R | S | R | S | I | 0.5 |
| S105 | *Escherichia coli* strain NRBC 102203 | S | S | S | S | R | I | S | R | I | S | 0.2 |
| S115 | *Enterobacter cloacae* strain DSM30054 13047 | S | R | R | R | R | R | S | R | R | I | 0.7 |
| S117 | *Klebsiella pneumoniae* strain TA6363 | S | R | R | S | R | R | R | R | R | I | 0.7 |
| S121 | *Pseudomonas aeruginosa* DSM 50071 | S | nd | nd | nd | nd | S | nd | nd | nd | S | 0 |
| S134 | *Pseudomonas alcaliphila* strain NRBC 102411 | S | nd | nd | nd | nd | S | nd | nd | nd | S | 0 |

CN= Gentamycin; SXT= Trimethoprim-sulfamethoxazole; TE= Tetracycline; FOX= Cefoxitin; AMC= Amoxicillin-clavulanic acid; CIP= Ciprofloxacin; STR= Streptomycin; CTX= Cefotaxime; CAZ= Ceftazidime; IMP= Imipenem; MAR= Multiple Antibiotic Resistance; I= intermediate; R= Resistance; S= Susceptible; nd= non-determinate

Multidrug resistance (MDR) were found in 54.2% of the bacteria in accordance with standard protocol (25). *Acinetobacter baumannii* and all isolates of *Enterobacter* spp. and 71.4% of *K. pneumoniae* were MDR bacteria.

Multiple antibiotic resistance (MAR) index ≥0.3 is present in 15 of the isolates (Table 3). Rarely pathogenic bacteria such as *A. soli, A. junii* and *P. alcaliphila* which were never diagnosed in LRTI or human infection have low MAR Index (Table 3). However, *P. parafulva* (S89) had MAR index of 0.3 (Table 3). *Enterobacter cloacae* and *Enterobacter hormaechei* subsp *xianfangensis* have very high MAR index and are more prevalent in the population (Table 3). Other common opportunistic and nosocomial pathogens such as *K. pneumoniae* and *A. baumannii* also have MAR Index within 0.3 to 0.8 (Table 3). Some bacteria of the same species but from different patients have different MAR index: *Klebsiella Pneumoniae* (is found in 7 patients and have varying MAR index (Table 3). Surprisingly, *P. aeruginosa* was found in only a patient with 100% susceptibility to all the antibiotics of choice that were tested. Hence a zero MAR index (Table 3).

### Antibiotics resistance and susceptibility correlation pattern

Antibiotic resistance correlation and regression analysis showed that bacterial resistance to one antibiotic correlates with resistance to other antibiotics and MAR index (Table 4). for example, resistance to CN correlates with CIP (p= 0.0008; r=0.54) and negatively with MAR index (p=0.003, r = −0.60). Only the CN, CIP and IPM show negative correlation with the MAR index while AMC and CTX show a positive correlation (Table 4). STX resistance and susceptibility pattern correlates more with those of other antibiotics (Table 4). CN and IPM are the two most effective antibiotics among this population (Table 4).

**Table 4:**
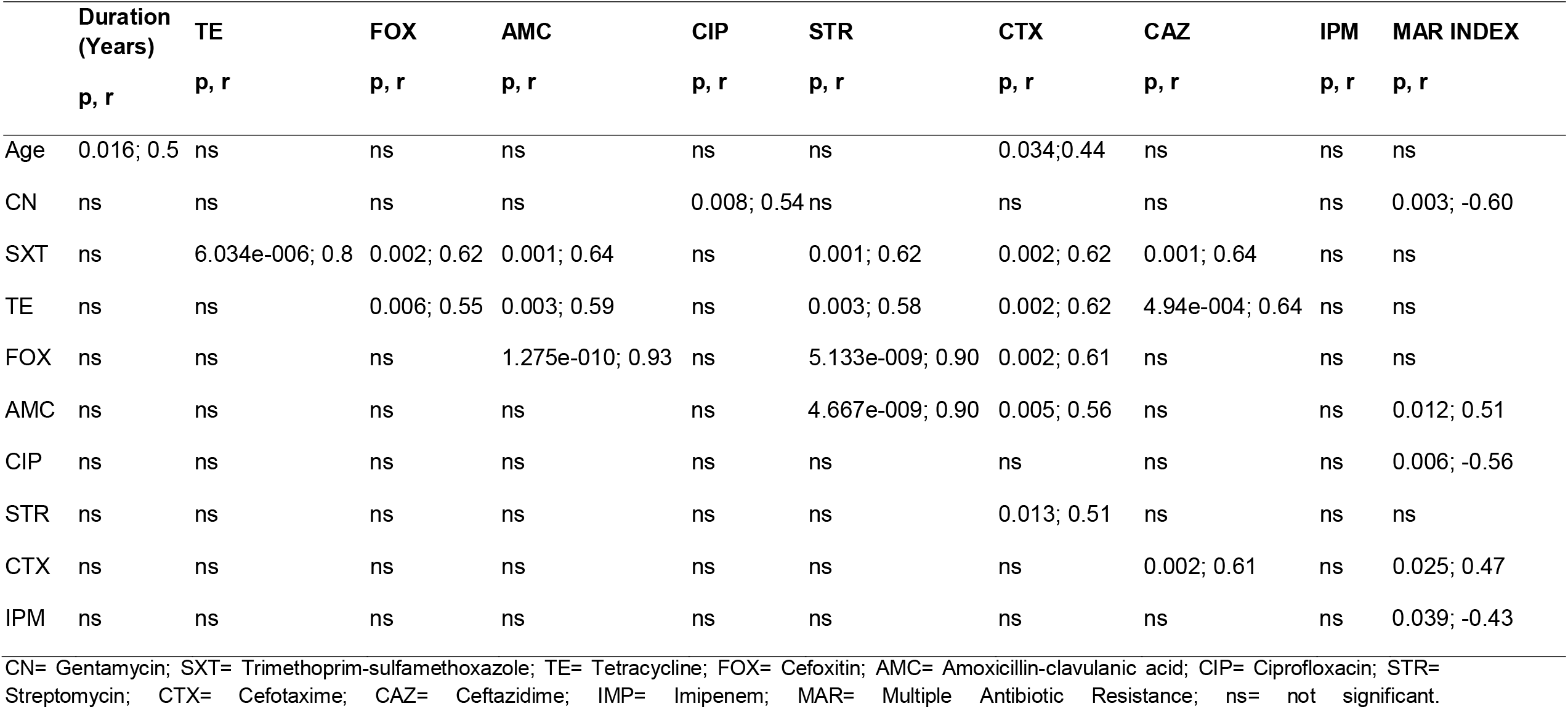
Antibiotics resistance correlation and regression pattern

|  | **Duration**<br>**(Years)** | **TE** | **FOX** | **AMC** | **CIP** | **STR** | **CTX** | **CAZ** | **IPM** | **MAR INDEX** |
| --- | --- | --- | --- | --- | --- | --- | --- | --- | --- | --- |
|  | **p, r** | **p, r** | **p, r** | **p, r** | **p, r** | **p, r** | **p, r** | **p, r** | **p, r** | **p, r** |
| Age | 0.016; 0.5 | ns | ns | ns | ns | ns | 0.034; 0.44 | ns | ns | ns |
| CN | ns | ns | ns | ns | 0.008; 0.54 | ns | ns | ns | ns | 0.003; -0.60 |
| SXT | ns | 6.034e-006; 0.8 | 0.002; 0.62 | 0.001; 0.64 | ns | 0.001; 0.62 | 0.002; 0.62 | 0.001; 0.64 | ns | ns |
| TE | ns | ns | 0.006; 0.55 | 0.003; 0.59 | ns | 0.003; 0.58 | 0.002; 0.62 | 4.94e-004; 0.64 | ns | ns |
| FOX | ns | ns | ns | 1.275e-010; 0.93 | ns | 5.133e-009; 0.90 | 0.002; 0.61 | ns | ns | ns |
| AMC | ns | ns | ns | ns | ns | 4.667e-009; 0.90 | 0.005; 0.56 | ns | ns | 0.012; 0.51 |
| CIP | ns | ns | ns | ns | ns | ns | ns | ns | ns | 0.006; -0.56 |
| STR | ns | ns | ns | ns | ns | ns | 0.013; 0.51 | ns | ns | ns |
| CTX | ns | ns | ns | ns | ns | ns | ns | 0.002; 0.61 | ns | 0.025; 0.47 |
| IPM | ns | ns | ns | ns | ns | ns | ns | ns | ns | 0.039; -0.43 |

CN= Gentamycin; SXT= Trimethoprim-sulfamethoxazole; TE= Tetracycline; FOX= Cefoxitin; AMC= Amoxicillin-clavulanic acid; CIP= Ciprofloxacin; STR= Streptomycin; CTX= Cefotaxime; CAZ= Ceftazidime; IMP= Imipenem; MAR= Multiple Antibiotic Resistance; ns= not significant.

**Table 5:**
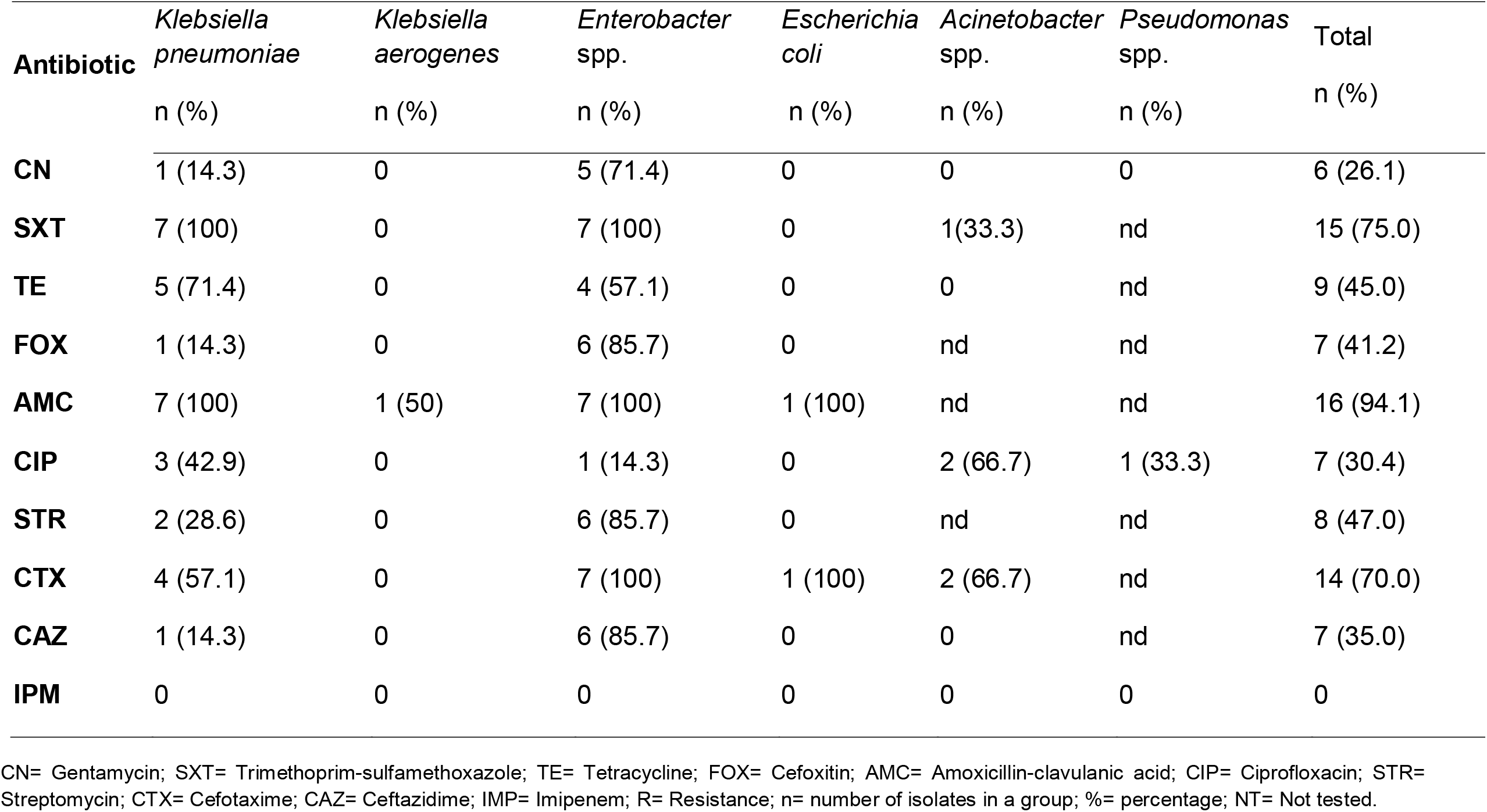
Antibiotic Resistance Profile of Bacterial Isolates

| **Antibiotic** | *Klebsiella pneumoniae* | *Klebsiella aerogenes* | *Enterobacter* spp. | *Escherichia coli* | *Acinetobacter* spp. | *Pseudomonas* spp. | Total |
| --- | --- | --- | --- | --- | --- | --- | --- |
|  | n (%) | n (%) | n (%) | n (%) | n (%) | n (%) | n (%) |
| **CN** | 1 (14.3) | 0 | 5 (71.4) | 0 | 0 | 0 | 6 (26.1) |
| **SXT** | 7 (100) | 0 | 7 (100) | 0 | 1(33.3) | nd | 15 (75.0) |
| **TE** | 5 (71.4) | 0 | 4 (57.1) | 0 | 0 | nd | 9 (45.0) |
| **FOX** | 1 (14.3) | 0 | 6 (85.7) | 0 | nd | nd | 7 (41.2) |
| **AMC** | 7 (100) | 1 (50) | 7 (100) | 1 (100) | nd | nd | 16 (94.1) |
| **CIP** | 3 (42.9) | 0 | 1 (14.3) | 0 | 2 (66.7) | 1 (33.3) | 7 (30.4) |
| **STR** | 2 (28.6) | 0 | 6 (85.7) | 0 | nd | nd | 8 (47.0) |
| **CTX** | 4 (57.1) | 0 | 7 (100) | 1 (100) | 2 (66.7) | nd | 14 (70.0) |
| **CAZ** | 1 (14.3) | 0 | 6 (85.7) | 0 | 0 | nd | 7 (35.0) |
| **IPM** | 0 | 0 | 0 | 0 | 0 | 0 | 0 |

CN= Gentamycin; SXT= Trimethoprim-sulfamethoxazole; TE= Tetracycline; FOX= Cefoxitin; AMC= Amoxicillin-clavulanic acid; CIP= Ciprofloxacin; STR= Streptomycin; CTX= Cefotaxime; CAZ= Ceftazidime; IMP= Imipenem; R= Resistance; n= number of isolates in a group; %= percentage; NT= Not tested.

**Table 6:** Percentage Multi-Drug Antibiotic Resistance (MDR) Pattern of Bacteria from LRTI of HIV/AIDS Patients

| Bacteria | No. of antibiotic classes isolates were resistant to |  |  |  | Resistance pattern |
| --- | --- | --- | --- | --- | --- |
|  | Three<br>n (%) | Four<br>n (%) | Five<br>n (%) | Six<br>n (%) | MDR<br>n (%) |
| *Klebsiella pneumoniae*(n=7) | 1 (14.3) | 0 | 4 (57.1) | 0 | 5 (71.4%) |
| *Enterobacter cloacae* (n=4) | 0 | 0 | 2 (50.0) | 2 (50.0) | 4 (100.0%) |
| *E. hormaechei* subsp. *xianfangensis*(n=3) | 0 | 1 (33.3) | 1 (33.3) | 1 (33.4) | 3 (100.0%) |
| *Acinetobacter baumannii* (n=1) | 1 (100.0) | 0 | 0 | 0 | 1 (100.0%) |

MDR= multiple drug resistance; n= number; %= percentage. MDR was analysed based on the percentage number of antibiotics categories to which an organism was resistant. Percentage organisms that showed resistant to antibiotics in more than two antibiotic categories were classified as MDR.

**Table 7:** Prevalence of BLRTI with CD4 cell count

| HIV-Associated<br>Immunodeficiency | CD4 cell count<br>(cell/mm<sup>3</sup>) | number of<br>patients<br>recruited | No. of patients<br>with bacterial<br>infection<br>n (%) |
| --- | --- | --- | --- |
| Severe | <200 | 11 | 2 (18.2) |
| Advanced | 200-349 | 27 | 7 (25.9) |
| Mild | 350-499 | 30 | 10 (33.3) |
| None | ≥500 | 51 | 5 (9.8) |
|  |  | 119 | 24 (20.2) |

CD4= cluster of differentiation; mm= millimetre; HIV= human immunodeficiency virus; n= number; %= percentage

## 4.0 DISCUSSION

Antibiotic resistance features at the top of the global public health agenda, with a call for action to reduce its threat and the impact on patients suffering from drug-resistant infections. However, the prioritisation of policy action, research, and the evaluation of progress towards curbing the threat of antimicrobial resistance (AMR) is dependent on a greater level of understanding of its burden both globally and at the national level.

Frequent use of antibiotics among the HIV/AIDs patient is important to prevent enormous threat of opportunistic infection from bacterial pathogens. Hence, emerging bacterial resistance has the potential to be devastating in HIV/AIDs prevalent region due to widespread of host immunocompromise and absence of strict regulation in antibiotic usage in developing countries.

In this study, bacteria were associated with LRTI in HIV/AIDs patients visiting University of Abuja Teaching Hospital with a prevalence rate of 20.2%, similar to previous study (26).

Bacteria not reported to have been previously associated with Lower Respiratory Tract Infection (LRTI) and rarely pathogenic bacteria such as *Acinetobacter soli*; *Enterobacter hormaechei* subsp *xianfangensis*; *Acinetobacter junii*; *Pseudomonas parafulva, Comamonas testosteroni* and *Psudomonas alcaliphila* which are of plants, environment, hospital and normal flora aetiology were found among these HIV/AIDS patients. *Comamonas testosteroni* however have been reported to be associated with pneumonia of an HIV/AIDS patient (27). Common causes of LRTI (2,10,26,28,29), identified in this study were *Klebsiella pneumoniae, Enterobacter cloacae, Acinetobacter baumannii, Pseudomonas aeruginosa* and *Escherichia coli*.

The study shows that the prevalence of Bacterial LRTI varies with the age, gender and, CD4 cell counts of the patients. The prevalence appeared to be higher in male than in female. In Africa, men have worse treatment outcomes than women (30), and may be linked to their poor adherence to treatment compared to women (31). More bacterial isolates were found in patients with CD4 count <500 cell/mm^3^, showing that HIV/AIDS associated immunodeficiency is associated with increased risk for opportunistic infections (32,33). The vulnerability of the immunocompromised to many opportunistic pathogens makes them a good metrics to monitoring the spread of antibiotics resistance and the need to mount surveillance on the aetiologies of bacterial resistance to antibiotic from a One-health approach.

Isolates were mostly resistant to AMC, SXT and CTX. Bacteria like *K. pneumoniae* and *Enterobacter* spp., the most prevalent isolates showed to be highly resistant to SXT, AMC and CTX, up to 100% resistance. The high degree of resistance to SXT among HIV/AIDS patients observed in this study as with other reports (10,34), may be due to long term use of Trimethoprim-sulfamethoxazole as prophylaxis for *Pneumocystis carni* pneumonia (PCP) (now known as *Pneumocystis jirovenci* pneumonia) in accordance to WHO recommendation for HIV/AIDS patient (35,36). Studies have shown association between long-term use of Trimethoprim-sulfamethoxazole with increase in development of resistance to Trimethoprim-sulfamethoxazole among bacteria in HIV/AIDS patients (12,13). High degree of resistance to AMC may be due to an intrinsic resistance mechanism of *Enterobacter* spp. to β-lactamase inhibitors such as AMC (25), while that of CTX may be due to increasing widespread resistance of *enterobacteriaceae* to 3^rd^ generation cephalosporins (37).

Classification of bacteria according to their resistance pattern showed that 54.2% of the isolates were multidrug resistant (MDR) and are observed in *A. baumannii*, all isolates of *Enterobacter* spp and 71.4% of *K. pneumoniae*, similar to previous study (29). The occurrence of MDR isolates in HIV/AIDS patients, particularly, the *Enterobacteriaceae* associated with respiratory tract infection are becoming widespread (38–40).

HIV/AIDS is a risk factor for the emergence of antibiotic resistance (41). The practice of frequent and prolonged use of antibiotics for treatment and or prophylaxis due to increased risk to opportunistic infections because of AIDS-immunodeficiency is a potential for the development of multidrug-resistant bacteria due to selective pressure.

The emergence of Multiple Antibiotics Resistant (MAR) bacteria is a threat to human health (42). The emergence of MAR bacteria is due to indiscriminate use of antibiotics in clinical medicine, agriculture and aquaculture industries (43,44). Furthermore, MAR index is a measure of the spread of bacterial resistance in an environment (45). MAR Index ≥0.3 show that bacteria are from environment with high antibiotic pressure while MAR index ≤0.2 is indicative of source with less antibiotic pressure (46). Hence, the high MAR index, from 0.300.8 observed in 62.5% of isolates in this study show that there is high antibiotic pressure among the HIV/AIDS population due to indiscriminate use of antibiotics In low resource settings, antibiogram along with MAR index serve an important epidemiological tool to monitor drug resistance.

The increased MAR index >0.2 in 62.5% in *P. parafulva, Enterobacter cloacae, Enterobacter cloacae, Enterobacter hormaechei* subsp *xianfangensis, K. pneumoniae*; and *A. baumannii* shows high exposure to antibiotics. Hence, increasing prevalence of HIV/AIDs in developing countries could contribute to growing antibiotic resistance challenge. Hospital acquired infection and antibiotic resistance remain a major threat to patients. A hospital-associated pathogen, *Enterobacter cloacae* and *Klebsiella pneumoniae* with high MAR index are indicative of higher exposure to antibiotics as expected from within a hospital environment.

A yet unknown hospital associated pathogen, *Enterobacter hormaechei* subsp *xianfangensis* species was found to be common in this population with high MAR index. These opportunistic nosocomial bacteria alongside with *A. baumannii, also* show to be multi-Drug resistant (MDR). While a lot of focus has been on MRSA, we propose that these *Enterobacter cloacae, Klebsiella pneumoniae, Enterobacter hormaechei* subsp *xianfangensis, and A. baumannii* could pose potential threat like MRSA, especially, among the immunocompromised and could play a role in resistance gene transfer within the environment.

Bacteria with similar MDR and MAR index could be indicative of the same route of infection and vice versa. The study also show that bacteria of the same species but from different patients have different MAR index depending on their exposure to antibiotics. For example, *Klebsiella Pneumonia* showed different MAR index, ranging from 0.2-0.7 in different subjects. This implies that patients with similar infection are unlikely to respond to antibiotics the same way. Furthermore, organism response to antibiotics could be dependent on previous antibiotic exposure within the environment, like human host. We proposed that the presence of the same species of bacteria in different patients and with a different MAR index indicates different aetiology rather than being acquired from the same source. Hence, different prognosis, in terms of response to antibiotics could be observed in patients with LRTI when caused by same organism, like the *Klebsiella Pneumonia*.

A well-known MDR bacterium, *P. aeruginosa*, a specie of considerable medical importance, recognized for its ubiquity, intrinsic antibiotic resistant mechanism and its association with serious illnesses such as nosocomial infection, ventilator associated pneumonia and various sepsis syndrome (47), was found in a patient and with 100% susceptibility to all the antibiotics used in this study, hence, a zero MAR index. The aetiology of this *P. aeruginosa* could be from environment with no or lesser exposure to antibiotics. Hence, the susceptibility for a widely acclaimed resistant organism. In addition, the presence of this 100% susceptible isolate likely to have come from a non-antibiotic exposed environment shows that HIV/AIDs patients could be a vector for different isolates from different environment. This is also validated by the presence of other rarely pathogenic and environmental bacteria, infrequently isolated in clinical practice such as *Comamonas testosteroni, P. parafulva* and *P. alcaliphila*.

A correlation and regression analysis indicated that the susceptibility profile of isolates to a given antibiotic could correlate with other antibiotics as well as the MAR index across the isolates. Hence, the mechanism of resistance of bacterium might affect more than one antibiotic. Antibiotics, CIP, CN and IPM were found to be very effective against most of the organism and showed a negative correlation to the MAR index across isolates unlike the AMC and the CTX with positive correlation with MAR index. We propose that antibiotics with significant negative correlation with the MAR index are less likely to induce widespread resistance within the environment. The somewhat effective CIP and highly effective CN and IPM, even against MDR isolates are useful alternative therapy to down regulate selective pressure on other antibiotics, similar to previous report (48).

We proposed that negative MAR index correlation indicates a good antibiotic to which resistance is unlikely to develop when used across isolates. For example, exposure of Isolates to CN is unlikely to lead to increasing MAR index as the susceptibility/resistance pattern of isolates to CN negatively correlated the MAR index across bacterial population.

In conclusion, the challenge of antibiotics is deepening each day and there might be the need for a personalised antibiotic profiling rather than to base treatment on previously known susceptibility of a given bacterial pathogen or causative agents. There is need for a specialised bacteria MAR index database which shows increase in the rates of resistance among these organisms thus making antimicrobial susceptibility surveillance and testing more crucial in selecting empiric regimen or definitive treatment (49). Several factor including inconsistent access to highly active antiretroviral therapy (50), may be responsible for increasing prevalence of HIV/AIDs in developing countries like Nigeria. This place developing nations in danger of increasing HIV/AIDS prevalence and the risk of emerging antibiotic resistance, which could be compounded by resistant strain circulation and increasing vulnerability to drug-resistant opportunistic infection (50,51), as observed in patients with LRTI. HIV and immunocompromised patients therefore present a very good metrics to understanding the diversity and depth of antibiotic resistance among bacterial isolates in clinical setting, particularly in developing countries.

## Data Availability

Data are deposited in Figshare.The 16srRNA gene sequence of bacteria isolates from lower respiratory tract infection is deposited in GenBank: Accession number MZ662083-MZ662100

https://doi.org/10.6084/m9.figshare.15035742

## Data Availability Statement

Data repository is figshare: https://doi.org/10.6084/m9.figshare.15035742. This project contains the following underlying data

- Data file 1. Cohort Table
- Data file 2. Biochemical identification of bacterial isolates
- Data file 3. Zone diameter of inhibition of antibiotic susceptibility test
- Data file 4. Antibiotic susceptibility test interpretation

The 16srRNA gene sequence of bacteria isolates from lower respiratory tract infection is deposited in GenBank: Accession number <u>MZ662083-MZ662100</u>. Data are available under the terms of the Creative Commons Zero “No rights reserved” data waiver (CC0 1.0 Public dedication).

## Authors contribution

JIO and SEA performed the conceptualisation methodology, investigation, formal analysis, data analysis and writing original draft and review. JIO and VGK collected the samples. SEA supervised the laboratory investigation and manuscript writing. VGK performed patient examination and establish the clinical diagnosis. ROB and OJA supervised JIO’s project and contributed to the review and editing of the manuscript.

SE-A, SS, and TS participated in the review and the editing of the manuscript. GEE supported the analysis of the data, data curation and proof-reading of the manuscript.

## Acknowledgement

The Authors would like to thank all the patients that gave their consent to participate in this study. We thank the laboratory staff of the University of Abuja Teaching hospital (UATH) that participated in supporting this work in one way or the other, particularly the friendly atmosphere within the laboratory. We thank Dr. Anthony Holder for the support and advise he gave during this work and allowing part of the work to be done in his laboratory at the Francis crick institute, London. The Authors thank the EBIHAS virtual research group for the contribution to the project and HolyNations International Ministries for the support in sponsoring this work, particularly the sample collection and the laboratory analysis and the stipends provided for the student. We thank the Department of Pharmaceutical Microbiology, Ahmadu Bello university, Zaira for their support during this work, which is part of MSc, dissertation. We thank Dr. Bolaji R.O and prof. Onaolapo J.A. for their supervisory role at the Ahmadu Bello university during the studies that led to this work. Thanks to Dr. Vivian Gga Kwaghe at the University of Abuja Teaching Hospital for supporting the work, particularly in clinical diagnosis and sample collection and the Health Research Ethics Committee (HREC) of the University of Abuja Teaching Hospital. Thanks to Dr. Samuel Abah for supporting to ensure we secure support from HolyNations International ministries for his skill and supervisory role during project design and actual laboratory analysis in the UK.

## References

1. Feikin DR, Feldman C, Schuchat A, Janoff EN. Global strategies to prevent bacterial pneumonia in adults with HIV disease [Internet]. Vol. 4, Lancet Infectious Diseases. Lancet Infect Dis; 2004 [cited 2021 Feb 5]. p. 445–55. Available from: https://pubmed.ncbi.nlm.nih.gov/15219555/

2. Ojha CR, Rijal N, Khagendra KC, Palpasa K, Kansakar P, Gupta BP, et al. Lower respiratory tract infections among HIV positive and control group in Nepal. VirusDisease. 2015;26(1–2):77–81.

3. Huson MAM, Hoogendijk AJ, De Vos AF, Grobusch MP, Van Poll T Der. The impact of HIV infection on blood leukocyte responsiveness to bacterial stimulation in asymptomatic patients and patients with bloodstream infection. J Int AIDS Soc. 2016;19(1).

4. Center for Disease Control and Prevention. Pneumocystis pneumonia -- Los Angeles 1981 [Internet]. Vol. 30. 1981 [cited 2021 Feb 9]. Available from: https://stacks.cdc.gov/view/cdc/50022

5. Benito N, Moreno A, Miro JM, Torres A. Pulmonary infections in HIV-infected patients: An update in the 21st century. Eur Respir J. 2012;39(3):730–45.

6. Morris A, Crothers K, Beck JM, Huang L. An official ATS workshop report: Emerging issues and current controversies in HIV-associated pulmonary diseases. Proc Am Thorac Soc. 2011;8(1):17–26.

7. Denegre AA, Myers K, Fefferman NH. Impact of strain competition on bacterial resistance in immunocompromised populations. Antibiotics. 2020;9(3).

8. Bratzler DW, Houck PM. Antimicrobial prophylaxis for surgery: An advisory statement from the national surgical infection prevention project [Internet]. Vol. 38, Clinical Infectious Diseases. Clin Infect Dis; 2004 [cited 2021 Feb 5]. p. 1706–15. Available from: https://pubmed.ncbi.nlm.nih.gov/15227616/

9. Laxminarayan R, Duse A, Wattal C, Zaidi AKM, Wertheim HFL, Sumpradit N, et al. Antibiotic resistance-the need for global solutions [Internet]. Vol. 13, The Lancet Infectious Diseases. Elsevier; 2013 [cited 2021 Feb 5]. p. 1057–98. Available from: http://www.thelancet.com/article/S1473309913703189/fulltext

10. O-B, Oluyege AO. Antibiotics Resistance of Bacteria Associated with Pneumonia in HIV / AIDS Patients in Nigeria. 2014;2(6):138–44.

11. Blair JMA, Webber MA, Baylay AJ, Ogbolu DO, Piddock LJV. Molecular mechanisms of antibiotic resistance. Nat Rev Microbiol [Internet]. 2015;13(1):42–51. Available from: http://dx.doi.org/10.1038/nrmicro3380

12. Martin, J.N., D.A. Rose, W.K. Hadley, F. Perdreau-Remington, P.K. Lam and JLG. Emergence of trimethoprim-sulfamethoxazole resistance in the AIDS era. Clin Infect Dis. 1999;180:1809–1818.

13. Zachariah R., Harries A.D., Spielmann, M.P., Arendt, V. Nchingula, D., Mwenda, .R., Courtielle, O. Kirpach, P., Mwale, B. and Salaniponi, F.M.L. Zachariah R., Harries A.D., Spielmann, M.P., Arendt, V. Nchingula, D., Mwenda, .R., Courtielle, O. Kirpach, P. FML. Changes in Escherichia coli resistance to cotrimoxazole in tuberculosis patients and in relation to cotrimoxazole prophylaxis in Thyolo, Malawi. Trans R Soc Trop Med Hyg. 2002;96:202–4.

14. World Health Organisation. Global health estimates 2016: Death by causes, ages, sex, by country and by region 2000-2016. https://www.who.int/healthinfo/global_burden_disease/estimates/en/. 2018. p. 1–3.

15. Morgan DJ, Okeke IN, Laxminarayan R, Perencevich EN, Weisenberg S. Non-prescription antimicrobial use worldwide: A systematic review [Internet]. Vol. 11, The Lancet Infectious Diseases. Elsevier; 2011 [cited 2021 Feb 5]. p. 692–701. Available from: http://www.thelancet.com/article/S1473309911700548/fulltext

16. Okeke IN, Lamikanra A, Edelman R. Socioeconomic and behavioral factors leading to acquired bacterial resistance to antibiotics in developing countries [Internet]. Vol. 5, Emerging Infectious Diseases. Centers for Disease Control and Prevention (CDC); 1999 [cited 2021 Feb 5]. p. 18–27. Available from: https://www.nc.cdc.gov/eid/article/5/1/99-0103_article

17. Gülsüm İclal Bayhan; Gönül Tanır; İbrahim Karaman; Şengül Özkan. Comamonas testosteroniu: An Unusual Bacteria Associated with Acute Appendicitis. Balkan Med J. 2013;30:447–8.

18. Tiwari S, Nanda M. Bacteremia caused by Comamonas testosteroni an unusual pathogen. J Lab Physicians [Internet]. 2019 Jan [cited 2021 Feb 10];11(01):087–90. Available from: /pmc/articles/PMC6437828/

19. Cheesbrough M. District Laboratory Practice in Tropical Countries. 2nd Editio. New York: Cambridge University Press; 2006. 62–76 p.

20. Cheesbrough M. District laboratory practice in tropical countries, second edition. District Laboratory Practice in Tropical Countries, Second Edition. 2006.

21. National Committee for Clinical Laboratory Standards N. M02-A12: Performance Standards for Antimicrobial Disk Susceptibility Tests; Approved Standard— Twelfth Edition. Clin Lab Stand Inst. 2015;

22. Clinical and Laboratory Standard Institutes. Performance standards for antimicrobial susceptibility testing. 27th ed. CLSI supplement M100. Wayne, PA: Clinical and Laboratory Standards Institute. Performance standards for antimicrobial susceptibility testing. 27th ed. CLSI supplement M100. Wayne, PA: Clinical and Laboratory Standards Institute. 2017. 282 p.

23. European Committee on Antimicrobial Susceptibility Testing. Breakpoint Tables for Interpretation of MICs and Zone Diameters. 2019;0–99. Available from: http://www.eucast.org.

24. Vashist Hemraj, Sharma Diksha GA. A Review on Commonly Used Biochemical Test for Bacteria. Innovare J Life Sci. 2013;1(1):221–30.

25. Magiorakos A, Srinivasan A, Carey RB, Carmeli Y, Falagas ME, Giske CG, et al. Bacteriau□: an International Expert Proposal for Interim Standard Definitions for Acquired Resistance. 2011;

26. Ramanlal Shah R, Professor A. Microbiological profile of respiratory tract infections among HIV-infected and HIV-non infected patients: A comparative study. 2016;3(2):51–9. Available from: http://iaimjournal.com/

27. Steinberg, J.P. and Burd EM. Other Gram Negative and Gram Variable Bacilli. In: Mandell, Douglas, and Bennett’s Principles and Practice of Infectious Diseases. 2015.

28. Khushbu Y SP. Bacteriological Profile of Lower Respiratory Tract Infection (LRTI) among HIV Seropositive Cases in Central Terai of Nepal. Int J Curr Microbiol Appl Sci. 2015;4(11):431–42.

29. Vishwanath S, Chawla K, Gopinathan A. Multidrug resistant Gram-negative bacilli in lower respiratory tract infections. 2013;5(4):323–7.

30. Druyts E, Dybul M, Kanters S, Nachega J, Birungi J, Ford N, et al. Male sex and the risk of mortality among individuals enrolled in antiretroviral therapy programs in Africa: A systematic review and meta-analysis. AIDS. 2013 Jan 28;27(3):417–25.

31. Hawkins C, Chalamilla G, Okuma J, Spiegelman D, Hertzmark E, Aris E, et al. Sex differences in antiretroviral treatment outcomes among HIV-infected adults in an urban Tanzanian setting. AIDS. 2011 Jun 1;25(9):1189–97.

32. Ledergerber B, Weber R, Hirschel B. AIDS-Related Opportunistic Illnesses. Library (Lond). 2009;8–12.

33. Jason M Brenchley, David A Price, Timothy W Schacker, Tedi E Asher, Guido Silvestri, Srinivas Rao, Zachary Kazzaz, Ethan Bornstein, Olivier Lambotte, Daniel Altmann, Bruce R Blazar, Benigno Rodriguez, Leia Teixeira-Johnson, Alan Landay, Jeffrey N Martin, SGD&DCD. Microbial translocation is a cause of systemic immune activation in chronic HIV infection. Nat Med [Internet]. 2016;12(12):1365–1371. Available from: https://doi.org/10.1038/nm1511

34. Kebede A, Aragie S, Shimelis T. The common enteric bacterial pathogens and their antimicrobial susceptibility pattern among HIV-infected individuals attending the antiretroviral therapy clinic of Hawassa university hospital, southern Ethiopia. Antimicrob Resist Infect Control. 2017;6(1):1–7.

35. World Health Organisation. Guidelines on co-trimoxazole prophylaxis for HIV-related infections among children, adolescents and adults: recommendations for a public health approach. Geneva 27, Switzerland. 2006. p. 9–10.

36. World Health Organisation. Guidelines on Post-Exposure Prophylaxis for Hiv and the Use of Co-Trimoxazole Prophylaxis for Hiv-Related Infections Among Adults, Adolescents and Children: Recommendations for a Public Health Approach. Recomm a public Heal approachDecember Suppl to Consol ARV Guidel WHO Geneva. 2014;(December):52.

37. Pitout, J.D. and Laupland KB. Extended-spectrum β-lactamase–producing Enterobacteriaceae: an emerging public-health concern. Lancet Infect Dis. 2008;8:159–66.

38. Aliberti, S., Di Pasquale, M., Zanaboni, A.M., Cosentini, R., Brambilla, A.M., Seghezzi, S., Tarsia, P., Mantero, M. and Blasi F. Stratifying risk factors for multidrug-resistant pathogens in hospitalized patients coming from the community with pneumonia. Clin Infect Dis. 2012;54:470–8.

39. Maruyama, T., Fujisawa, T., Okuno, M., Toyoshima, H., Tsutsui, K., Maeda, H., Yuda, H., Yoshida, M., Kobayashi, H. and Taguchi O. A new strategy for healthcare-associated pneumonia: a 2-year prospective multicenter cohort study using risk factors for multidrug-resistant pathogens to select initial empiric therapy. Clin Infect Dis. 2013;57:1373–83.

40. Khawcharoenporn T, Vasoo S, Singh K. Urinary Tract Infections due to Multidrug-Resistant Enterobacteriaceae: Prevalence and Risk Factors in a Chicago Emergency Department. Emerg Med Int. 2013;2013:1–7.

41. Denegre AA, Mbah MLN, Myers K, Id NHF. Emergence of antibiotic resistance in immunocompromised host populations□: A case study of emerging antibiotic resistant tuberculosis in AIDS patients. PLoS One [Internet]. 2019;14(2):1–13. Available from: http://dx.doi.org/10.1371/journal.pone.0212969

42. Gunst JJ, Langlois MR, Delanghe JR, De Buyzere ML, Leroux-Roels GG. Serum creatine kinase activity is not a reliable marker for muscle damage in conditions associated with low extracellular glutathione concentration. Clin Chem. 1998;

43. Al-Dulaimi MMK, Mutalib SA, Ghani MA, Zaini NAM, Ariffin AA. Multiple antibiotic resistance (Mar), plasmid profiles, and dna polymorphisms among vibrio vulnificus isolates. Antibiotics. 2019;8(2).

44. Tan CW, Malcolm TTH, Kuan CH, Thung TY, Chang WS, Loo YY, et al. Prevalence and antimicrobial susceptibility of Vibrio parahaemolyticus isolated from short mackerels (Rastrelliger brachysoma) in Malaysia. Front Microbiol. 2017;8(JUN).

45. Ehinmidu JO. Antibiotics susceptibility patterns of urine bacterial isolates in Zaria, Nigeria. Trop J Pharm Res. 2005;2(2):223–8.

46. Krumperman PH. Multiple Antibiotic Resistance Indexing of Escherichia coli to Identify High-Risk Sources of Fecal Contamination of Foodst [Internet]. Vol. 46, APPLIED AND ENVIRONMENTAL MICROBIOLOGY. 1983 [cited 2021 Feb 5]. Available from: http://aem.asm.org/

47. Osundiya O, Oladele R, Oduyebo O. Multiple Antibiotic Resistance (MAR) indices of Pseudomonas and Klebsiella species isolates in Lagos University Teaching Hospital. African J Clin Exp Microbiol. 2013 Aug;14(3).

48. WHO. WHO publishes World Malaria Report 2018. World Malar Rep. 2018;

49. Yousaf MZ, Zia S, Babar ME, Ashfaq UA. The epidemic of HIV/AIDS in developing countries; The current scenario in Pakistan. Vol. 8, Virology Journal. 2011.

50. DeNegre AA, Myers K, Fefferman NH. Impact of chemorophylaxis policy for AIDS-immunocompromised patients on emergence of bacterial resistance. PLoS One. 2020;15(1).

51. Laxminarayan R, Heymann DL. Challenges of drug resistance in the developing world. Vol. 344, BMJ (Online). 2012.

